# Development of a new trauma dataset over 38 years from the Young Finns Study

**DOI:** 10.64898/2026.08.26.26361417

**Authors:** Aino Saarinen, Timo Asikainen, Terho Lehtimäki, Olli Raitakari, Liisa Keltikangas-Järvinen

## Abstract

**Background:** Previous trauma research includes many limitations, such as the scarcity of pretraumatic health measurements and assessment of traumatic experiences with a broad scope across the lifespan. To respond to these gaps, we aimed to develop a new, prospective, population-based trauma dataset from childhood to middle age.

**Methods:** We used the Young Finns Study that is a population-based, multi-generational, prospective study (n = 3596 for the main generation). It has started in 1980 (baseline assessment) and includes follow-ups in 1983, 1986, 1989, 1992, 1997, 2001, 2007, 2011/2012, and 2018–2020. From the 38-year follow-up and ten measurement points of the YFS, we collected all relevant trauma variables, including both free-format and structured questions that both the participants and their parents responded to. By a data-driven case-to-case analysis, we developed a scale to numerically capture variation in the quality of the experiences.

**Results:** Our final dataset captured a total of 7769 traumatic experiences. We also developed the Traumatic Experience Severity Scale (TESS), including six subscales such as shamefulness, rarity, danger to life or health, effects on everyday life, human-made physical threat, and whether the target person was within or outside one’s household. We also preprocessed the dataset to be later easily interleaved with other psychological, cardiovascular, and epigenetic variables of the YFS.

**Conclusions:** We believe this new trauma dataset with thousands of experiences across the lifespan provides new opportunities to multidisciplinary, lifelong trauma research.

## 1. Introduction

While the association between traumatic experiences and adverse mental health outcomes is well-known, literature includes crucial gaps. First, most studies have adopted a post-traumatic-only approach without attention to pretraumatic measurements. A likely reason for this is that there has been a lack of datasets including both pre- and posttraumatic health assessments. This is a crucial limitation because there are single studies suggesting that, for example, as much as 87% of those with PTSD had already had psychiatric symptoms years before their traumatic experiences (Van Der Velden et al., 2023). Second, most studies have focused on PTSD, although a majority – even more than 70% – of the exposed individuals eventually develop PTSD (1) and more than half report positively oriented posttraumatic growth (2), such as increased self-acceptance, compassion, or social networking. Third, a majority of studies have had a comparatively narrow focus on traumatic experiences, investigating only traumas in a specific age (e.g., childhood) or only a specific type of trauma (e.g., combat war exposure). Thus, studies assessing traumatic experiences across the lifespan and with a broad scope have been scarce.

In addition, there has been a lack of measures to assess the severity of traumatic experiences. This is in clear contrast with other psychiatric symptoms since a variety of measures have been developed to examine the severity of depressive symptoms, psychotic symptoms, anxiety, sleep disturbances, or most other psychiatric symptoms. Accordingly, many previous studies have defined the severity of traumatic experiences simply as the number of traumatic experiences, although the reliability of this approach is limited by that single experiences can substantially differ from each other in their quality.

To respond to these gaps of the previous literature, we aimed to 1) develop a new trauma dataset with a broad scope of experiences and across the lifespan, and 2) develop a new scale to assess the severity of traumatic experiences. We used the Young Finns Study (YFS) (3, 4) that provides exceptional possibilities for this kind of project: it includes a 38-year follow-up with ten measurement points of traumatic experiences, assessed both with structured and free-format questions and including responses from two generations (i.e., the participants and their parents). Further the YFS also includes a variety of health measures in the fields of e.g. mental health, cardiovascular health, and epigenetic factors that can be later integrated with this trauma data.

## 2. Methods

### 2.1. Participants: The Young Finns Study

The participants came from the prospective Young Finns Study (YFS). The sampling was designed to include a population-based sample of Finnish non-institutionalized citizens (selected using the population register of the Social Insurance Institution). The original sample at the baseline measurement (1980) consisted of 3596 participants belonging to six age cohorts born in 1962, 1965, 1968, 1971, 1974, and 1977. Follow-ups have been conducted in 1983, 1986, 1989, 1992, 2001, 2007, 2011/2012, and 2018–2020. Participants were aged between 41 and 56 years in the most recent follow-up.

The study was carried out in accordance with the Declaration of Helsinki. Before participation, all the participants or their parents, in case of the participant was under 18 years old, provided informed consent after the nature of the procedures had been fully explained. The participants have the right to withdraw from the study at any time. Before data collection, the study design has been approved by the ethical committees of all the Finnish universities conducting the study (University of Turku, Tampere University, University of Helsinki, University of Oulu, University of Kuopio). Thus, the earliest ethical approvals have been received in the 1970s. The most recent approval has been received in 2017, i.e. before the most recent follow-up (ETMK:68/1801/2017). The design of the YFS is described more exactly elsewhere (Pahkala et al., 2026; Raitakari et al., 2008) .

### 2.2. Trauma variables across the lifespan

First, we selected all relevant data that could provide information on participants’ traumatic experiences. In summary, the data were gathered in two generations and over multiple measurement points:

1. We had data on traumatic experiences over **a 38-year follow-up** between the baseline measurement (1980) and eight follow-ups (1983, 1986, 1989, 1992, 2001, 2007, 2011, 2018). The timeline is depicted in **Figure 1**.
2. We used **two types of questions on** traumatic experiences. First, we had structured **questions** on a specific life event (e.g., “Have you encountered a death of a friend?”) that were responded with yes/no and the year of that experience (e.g., “in 2006”). Second, we had **open questions** that included a free-text field to report significant life events or catastrophes that had occurred in their life. The open questions also included an additional inquiry about the year of that experience.
3. We had data from **two generations** including participants (generation “G1”) and their parents (generation “G0”). As a main rule, parents responded to the questions when the participants were children. Additionally, in the follow-up of 2018, questions on “catastrophic experiences” were presented to both participants and parents, and many parents reported events/experiences that had occurred to their children.

**Figure 1.**
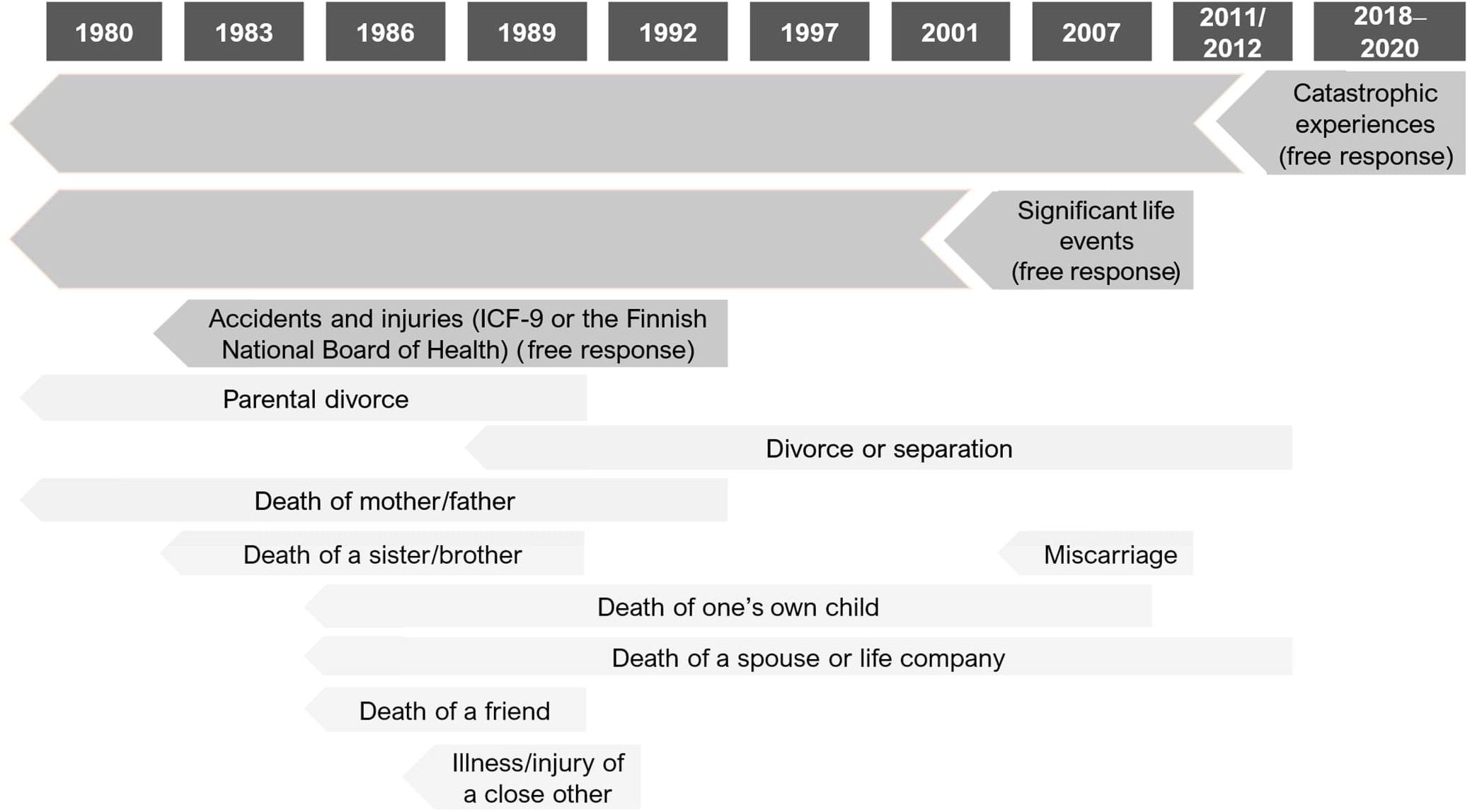
Timeline of the data collection on traumatic experiences.

**Table 1.** Measurement years and response scales of the trauma variables. Note: The frequencies presented before removal of duplicates between different follow-ups.

|  | Measurement year(s) | Response scale | Respondents: Participants (G1) or their parents (G0) | Frequency |
| --- | --- | --- | --- | --- |
| Death of mother or father | 1980, 1983, 1986, 1989, 1992 | Structured:<br>Yes/no and year | G0, G1 | 743 |
| Parental divorce | 1980, 1983, 1986, 1989 | Structured:<br>Yes/no and year | G0, G1 | 636 |
| Death of a sister/brother | 1983, 1986, 1989 | Structured:<br>Yes/no and year<br>Possible to report several deaths | G0, G1 | 104 |
| Severe illness or injury of a friend or a close other | 1989 | Structured:<br>Yes/no and year | G1 | 264 |
| Death of a friend | 1986, 1989 | Structured:<br>Yes/no and year | G1 | 260 |
| Divorce/separation (one's own partner) | 1989, 1992, 2001, 2007, 2011 | Structured:<br>Yes/no and year<br>Possible to report several divorces in 2001 and 2007 | G1 | 765 |
| Death of life company | 1986, 1989 | Structured:<br>Yes/no and year | G1 | 14 |
| Death of spouse | 1992, 2001, 2007, 2011 | Structured:<br>Yes/no and year | G1 | 79 |
| Miscarriage | 2007 | Structured:<br>Yes/no and year<br>Possible to report several miscarriages | G1 | 483 |
| Death of one's own child | 1986, 1989, 1992, 2001, 2007 | Structured:<br>Yes/no and year<br>Possible to report several deaths | G1 | 98 |
| Accidents and injuries that required medical treatment | 1983, 1986, 1989, 1992 | Free response:<br>At most 4 events could be reported; those were encoded in line with the ICF-9 or the Finnish National Board of Health<br>Further details in **Tables 2 and 3** | G0, G1 | 2988 |
| Significant negative life event | 2007 | Free response:<br>Participants could utilize at most 4 free-text fields<br>Further details in **Table 4** | G1 | 936 |
| Catastrophic experience | 2018 | Free response:<br>Participants (G1) could utilize at most 4 free-text fields and their parents (G0) could utilize at most 3 free-text fields<br>Further details in **Table 5** | G0, G1 | 494 |
|  |  |  |  | **Total:**<br>**7864** |

**Table 2.**
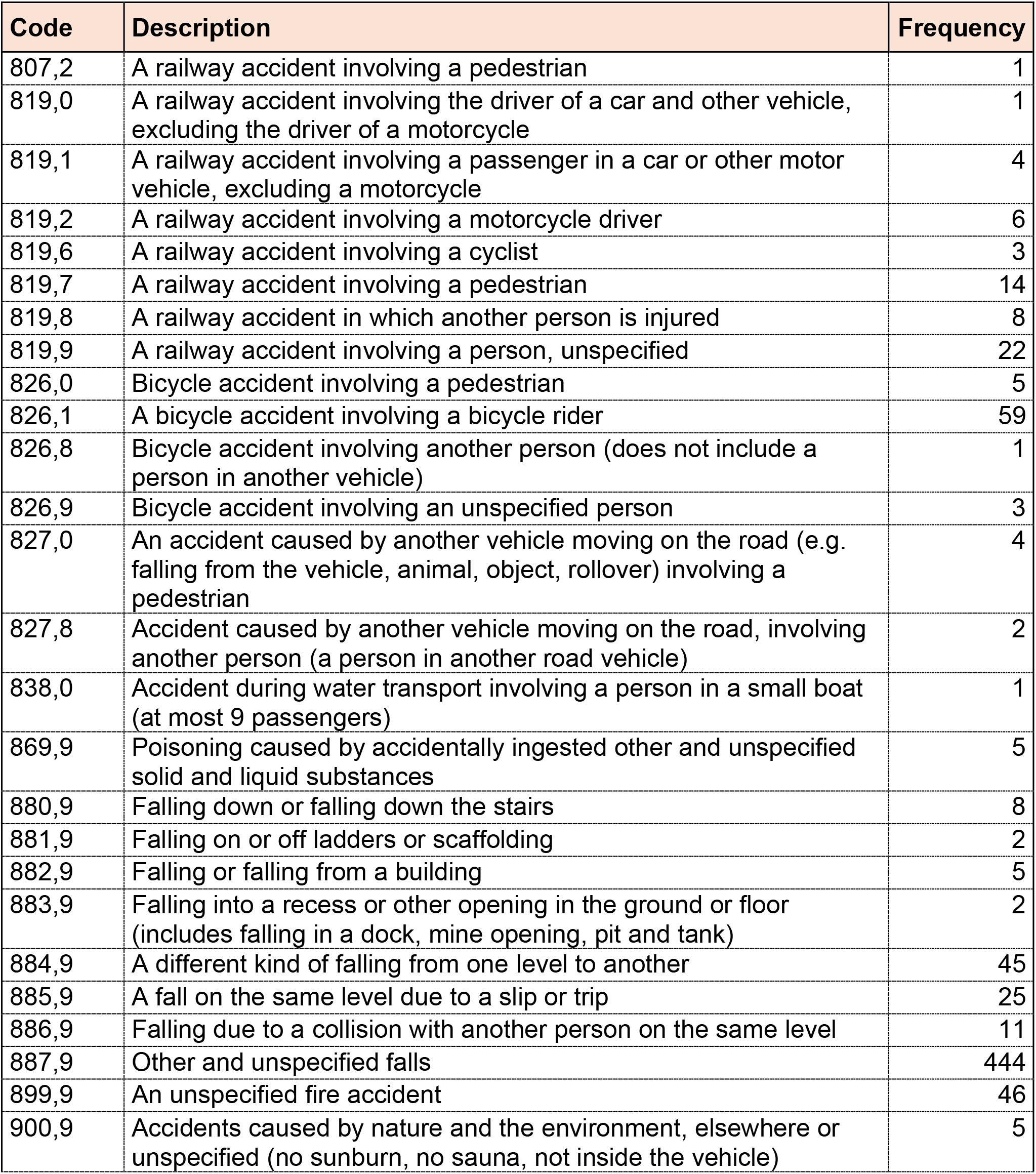
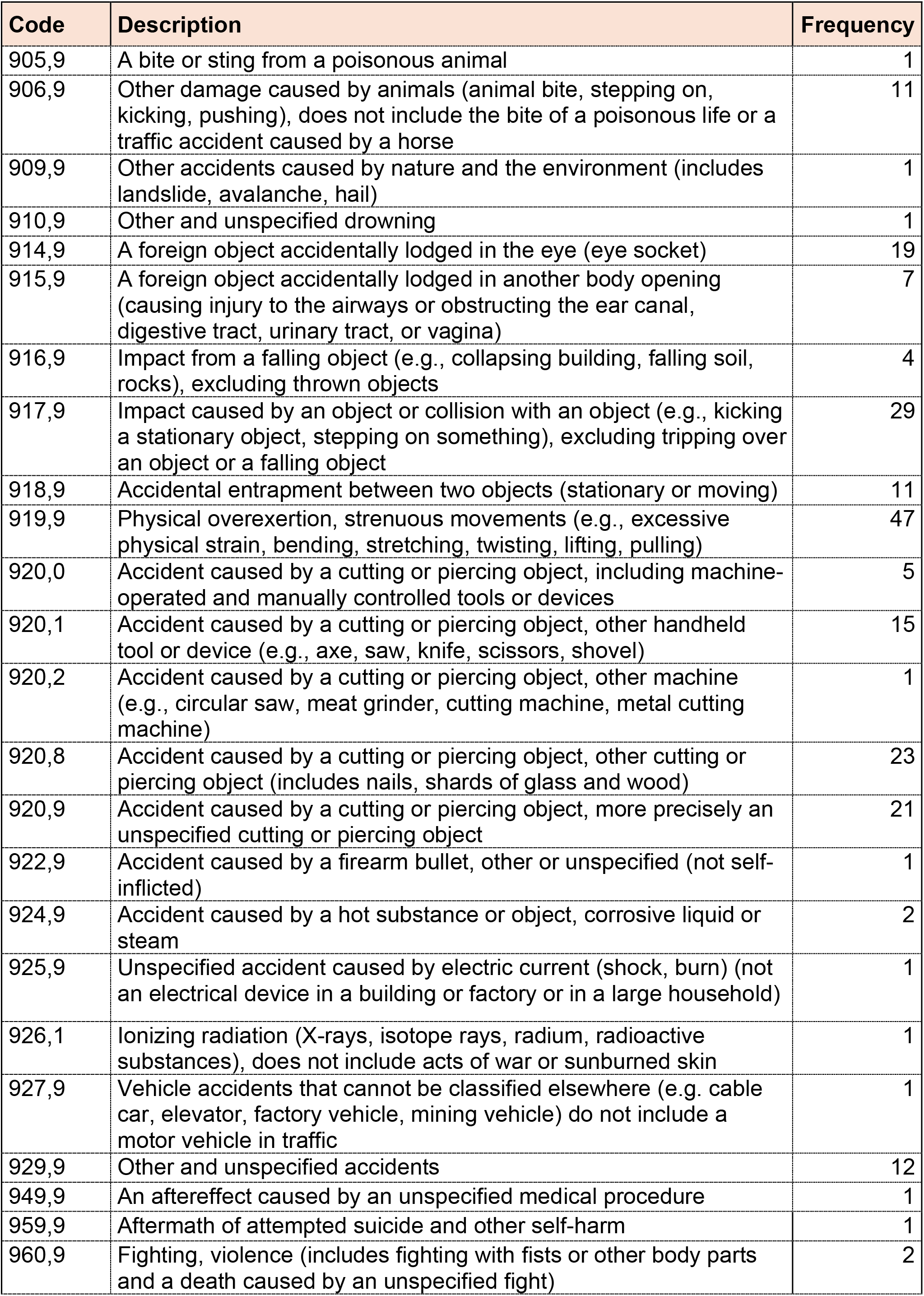
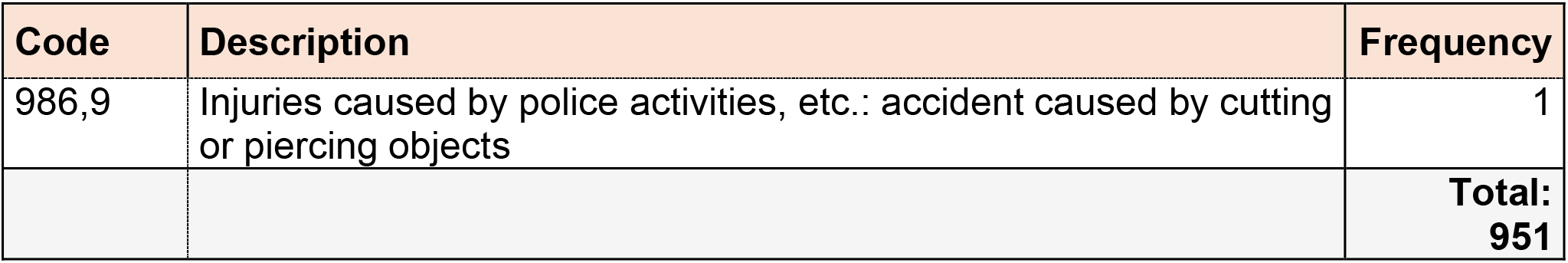
Classification of external causes of injury (the measurement of 1983 in the YFS) by the Finnish National Board of Health. Derived from https://www.julkari.fi/bitstream/handle/10024/135324/ICD8.pdf?sequence=1&isAllowed=y Note: This table includes only those codes for accidents/injuries that had occurred to the participants. The frequencies presented before removal of duplicates between different follow-ups.

| Code | Description | Frequency |
| --- | --- | --- |
| 807,2 | A railway accident involving a pedestrian | 1 |
| 819,0 | A railway accident involving the driver of a car and other vehicle, excluding the driver of a motorcycle | 1 |
| 819,1 | A railway accident involving a passenger in a car or other motor vehicle, excluding a motorcycle | 4 |
| 819,2 | A railway accident involving a motorcycle driver | 6 |
| 819,6 | A railway accident involving a cyclist | 3 |
| 819,7 | A railway accident involving a pedestrian | 14 |
| 819,8 | A railway accident in which another person is injured | 8 |
| 819,9 | A railway accident involving a person, unspecified | 22 |
| 826,0 | Bicycle accident involving a pedestrian | 5 |
| 826,1 | A bicycle accident involving a bicycle rider | 59 |
| 826,8 | Bicycle accident involving another person (does not include a person in another vehicle) | 1 |
| 826,9 | Bicycle accident involving an unspecified person | 3 |
| 827,0 | An accident caused by another vehicle moving on the road (e.g. falling from the vehicle, animal, object, rollover) involving a pedestrian | 4 |
| 827,8 | Accident caused by another vehicle moving on the road, involving another person (a person in another road vehicle) | 2 |
| 838,0 | Accident during water transport involving a person in a small boat (at most 9 passengers) | 1 |
| 869,9 | Poisoning caused by accidentally ingested other and unspecified solid and liquid substances | 5 |
| 880,9 | Falling down or falling down the stairs | 8 |
| 881,9 | Falling on or off ladders or scaffolding | 2 |
| 882,9 | Falling or falling from a building | 5 |
| 883,9 | Falling into a recess or other opening in the ground or floor (includes falling in a dock, mine opening, pit and tank) | 2 |
| 884,9 | A different kind of falling from one level to another | 45 |
| 885,9 | A fall on the same level due to a slip or trip | 25 |
| 886,9 | Falling due to a collision with another person on the same level | 11 |
| 887,9 | Other and unspecified falls | 444 |
| 899,9 | An unspecified fire accident | 46 |
| 900,9 | Accidents caused by nature and the environment, elsewhere or unspecified (no sunburn, no sauna, not inside the vehicle) | 5 |
| 905,9 | A bite or sting from a poisonous animal | 1 |
| 906,9 | Other damage caused by animals (animal bite, stepping on, kicking, pushing), does not include the bite of a poisonous life or a traffic accident caused by a horse | 11 |
| 909,9 | Other accidents caused by nature and the environment (includes landslide, avalanche, hail) | 1 |
| 910,9 | Other and unspecified drowning | 1 |
| 914,9 | A foreign object accidentally lodged in the eye (eye socket) | 19 |
| 915,9 | A foreign object accidentally lodged in another body opening (causing injury to the airways or obstructing the ear canal, digestive tract, urinary tract, or vagina) | 7 |
| 916,9 | Impact from a falling object (e.g., collapsing building, falling soil, rocks), excluding thrown objects | 4 |
| 917,9 | Impact caused by an object or collision with an object (e.g., kicking a stationary object, stepping on something), excluding tripping over an object or a falling object | 29 |
| 918,9 | Accidental entrapment between two objects (stationary or moving) | 11 |
| 919,9 | Physical overexertion, strenuous movements (e.g., excessive physical strain, bending, stretching, twisting, lifting, pulling) | 47 |
| 920,0 | Accident caused by a cutting or piercing object, including machine-operated and manually controlled tools or devices | 5 |
| 920,1 | Accident caused by a cutting or piercing object, other handheld tool or device (e.g., axe, saw, knife, scissors, shovel) | 15 |
| 920,2 | Accident caused by a cutting or piercing object, other machine (e.g., circular saw, meat grinder, cutting machine, metal cutting machine) | 1 |
| 920,8 | Accident caused by a cutting or piercing object, other cutting or piercing object (includes nails, shards of glass and wood) | 23 |
| 920,9 | Accident caused by a cutting or piercing object, more precisely an unspecified cutting or piercing object | 21 |
| 922,9 | Accident caused by a firearm bullet, other or unspecified (not self-inflicted) | 1 |
| 924,9 | Accident caused by a hot substance or object, corrosive liquid or steam | 2 |
| 925,9 | Unspecified accident caused by electric current (shock, burn) (not an electrical device in a building or factory or in a large household) | 1 |
| 926,1 | Ionizing radiation (X-rays, isotope rays, radium, radioactive substances), does not include acts of war or sunburned skin | 1 |
| 927,9 | Vehicle accidents that cannot be classified elsewhere (e.g. cable car, elevator, factory vehicle, mining vehicle) do not include a motor vehicle in traffic | 1 |
| 929,9 | Other and unspecified accidents | 12 |
| 949,9 | An aftereffect caused by an unspecified medical procedure | 1 |
| 959,9 | Aftermath of attempted suicide and other self-harm | 1 |
| 960,9 | Fighting, violence (includes fighting with fists or other body parts and a death caused by an unspecified fight) | 2 |
| 986,9 | Injuries caused by police activities, etc.: accident caused by cutting or piercing objects | 1 |
|  |  | **Total:**<br>**951** |

**Table 3.** List of the International Classification of Functioning, Disability and Health (ICD-9) E codes: external causes of injury (concerning the follow-ups of 1986, 1989, and 1992 in the YFS). Note: This table includes only those codes for accidents/injuries that had occurred to the participants. The frequencies presented before removal of duplicates between different follow-ups.

| Code | Description | Frequency |
| --- | --- | --- |
| E817,9 | Noncollision motor vehicle traffic accident while boarding or alighting injuring unspecified person | 1 |
| E818,9 | Other noncollision motor vehicle traffic accident injuring unspecified person | 2 |
| E819 | Motor vehicle traffic accident of unspecified nature | 1 |
| E819,0 | Motor vehicle traffic accident of unspecified nature injuring driver of motor vehicle other than motorcycle | 19 |
| E819,1 | Motor vehicle traffic accident of unspecified nature injuring passenger in motor vehicle other than motorcycle | 20 |
| E819,2 | Motor vehicle traffic accident of unspecified nature injuring motorcyclist | 28 |
| E819,6 | Motor vehicle traffic accident of unspecified nature injuring pedal cyclist | 15 |
| E819,7 | Motor vehicle traffic accident of unspecified nature injuring pedestrian | 6 |
| E819,8 | Motor vehicle traffic accident of unspecified nature injuring other specified person | 5 |
| E819,9 | Motor vehicle traffic accident of unspecified nature injuring unspecified person | 86 |
| E825,9 | Other motor vehicle nontraffic accident of other and unspecified nature | 5 |
| E826 | Pedal cycle accident | 1 |
| E826,0 | Pedal cycle accident injuring pedestrian | 1 |
| E826,1 | Pedal cycle accident injuring pedal cyclist | 60 |
| E826,9 | Pedal cycle accident injuring unspecified person | 8 |
| E827,0 | Animal-drawn vehicle accident injuring pedestrian | 3 |
| E827,8 | Animal-drawn vehicle accident injuring other specified person | 1 |
| E838,0 | Accident to watercraft causing submersion injuring occupant of small boat unpowered | 1 |
| E858,9 | Accidental poisoning by unspecified drug | 1 |
| E862,1 | Accidental poisoning by petroleum fuels and cleaners | 1 |
| E880,9 | Fall on same level from slipping, tripping or stumbling | 10 |
| E881,9 | Accidental fall on or from ladders or scaffolding | 2 |
| E882,8 | Accidental fall from or out of building or other structure | 1 |
| E882,9 | Fall on same level from slipping, tripping or stumbling | 5 |
| E883,9 | Accidental fall into other hole or other opening in surface | 1 |
| E884,9 | Other accidental fall from one level to another | 34 |
| E885,9 | Fall on same level from slipping, tripping or stumbling | 40 |
| E886,9 | Other and unspecified accidental falls on same level from collision pushing or shoving by or with other person | 6 |
| E887,9 | Fracture cause unspecified | 526 |
| E888,9 | Unspecified accidental fall | 1 |
| E889,5 | Accidental falls | 1 |
| E889,9 | Unintentional injury | 1 |
| E891,0 | Explosion caused by conflagration in other and unspecified building or structure | 1 |
| E895,9 | Accident caused by controlled fire in private dwelling | 1 |
| E898,9 | Accident caused by other specified fire and flames | 1 |
| E899,9 | Accident caused by unspecified fire | 9 |
| E900,9 | Accidents due to excessive heat of unspecified origin | 1 |
| E901,9 | Accident due to excessive cold of unspecified origin | 1 |
| E905,9 | Poisoning and toxic reactions caused by unspecified animals and plants | 2 |
| E906,9 | Unspecified injury caused by animal | 33 |
| E910,0 | Accidental drowning and submersion | 3 |
| E910,9 | Unspecified accidental drowning or submersion | 1 |
| E911,9 | Inhalation and ingestion of food causing obstruction of respiratory tract or suffocation | 1 |
| E912,9 | Inhalation and ingestion of other object causing obstruction of respiratory tract or suffocation | 1 |
| E914,9 | Foreign body accidentally entering eye and adnexa | 24 |
| E915,9 | Foreign body accidentally entering other orifice | 4 |
| E916,9 | Struck accidentally by falling object | 4 |
| E917,9 | Other accident caused by striking against or being struck accidentally by objects or persons with/without subsequent fall | 77 |
| E918,9 | Caught accidentally in or between objects | 26 |
| E919,5 | Accidents caused by prime movers except electrical motors | 1 |
| E919,6 | Accidents caused by transmission machinery | 1 |
| E919,9 | Accidents caused by unspecified machinery | 396 |
| E920,0 | Accidents caused by powered lawn mower | 13 |
| E920,1 | Accidents caused by other powered hand tools | 37 |
| E920,2 | Accidents caused by powered household appliances and implements | 6 |
| E920,8 | Accidents caused by other specified cutting and piercing instruments or objects | 44 |
| E920,9 | Accidents caused by unspecified cutting and piercing instrument or object | 73 |
| E922,9 | Accident caused by unspecified firearm missile | 3 |
| E924,9 | Accident caused by unspecified hot substance or object | 15 |
| E926,0 | Exposure to radiofrequency radiation | 1 |
| E928,8 | Other accidents | 1 |
| E928,9 | Unspecified accident | 2 |
| E929,9 | Late effects of unspecified accident | 292 |
| E929E | Late effects of accidental injury | 1 |
| E940,9 | Unspecified central nervous system stimulant causing adverse effects in therapeutic use | 18 |
| E943,9 | Unspecified agent primarily affecting the gastrointestinal system causing adverse effects in therapeutic use | 21 |
| E946,9 | Unspecified agent primarily affecting skin and mucous membrane causing adverse effects in therapeutic use | 5 |
| E949,9 | Other and unspecified vaccines and biological substances causing adverse effects in therapeutic use | 3 |
| E959,9 | Late effects of self-inflicted injury | 2 |
| E960,9 | Homicide and injury purposely inflicted by other persons: fight brawl rape | 16 |
| E969,9 | Late effects of injury purposely inflicted by other person | 1 |
| E989,9 | Late effects of injury undetermined whether accidentally or purposely inflicted | 1 |
| E994,9 | Injury due to war operations by unspecified destruction of aircraft | 1 |
|  |  | **Total:**<br>**2037** |

**Table 4.**
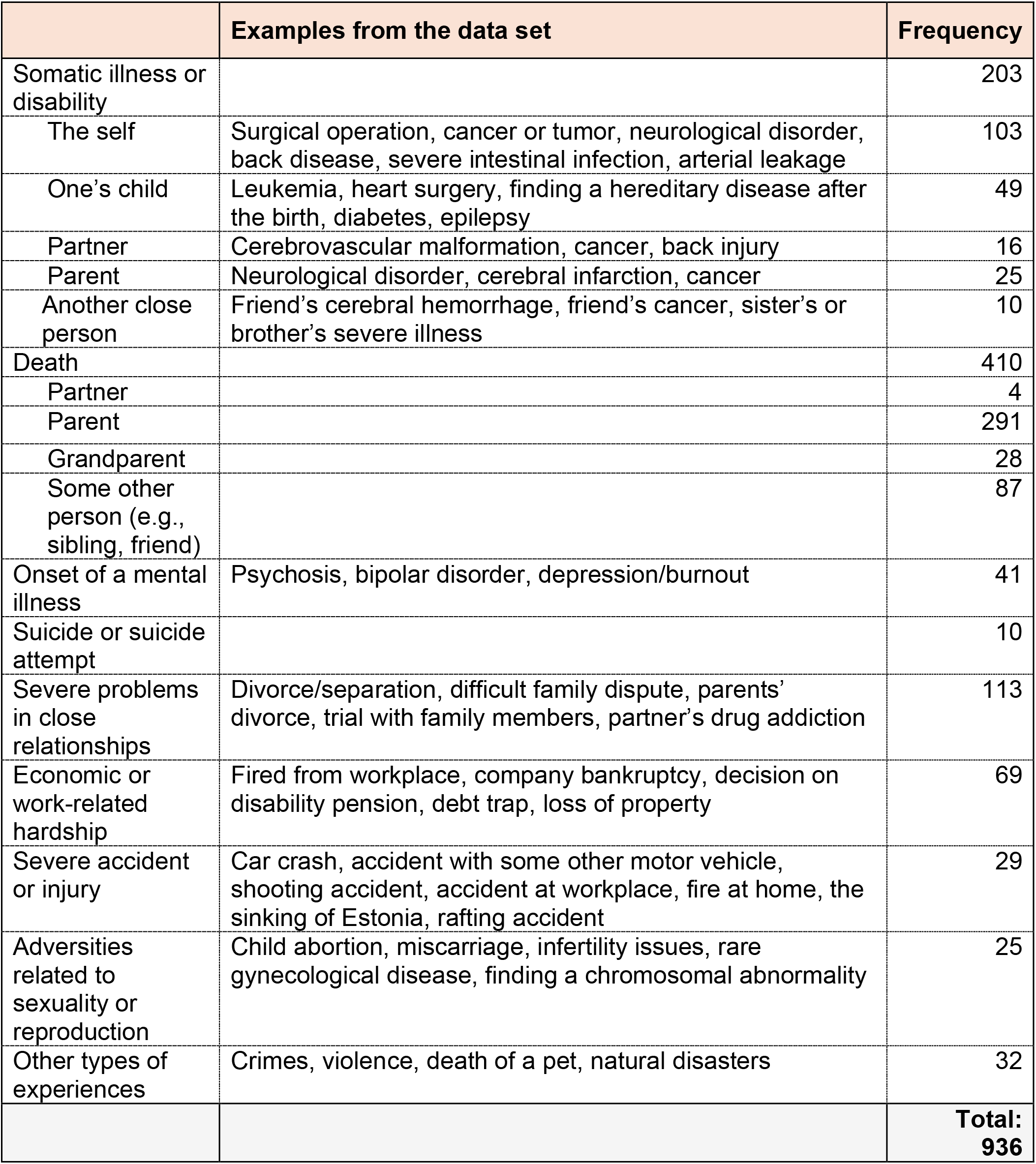
A summary of types of experiences that the participants reported in 2007 as significant negative life events. Note: The frequencies presented before removal of duplicates between different follow-ups.

**Table 5.** A summary of sorts of experiences that the YFS participants reported in 2018 as catastrophic life experiences. Note: The frequencies presented before removal of duplicates between different follow-ups.

|  | Examples from the YFS data set | Frequency |
| --- | --- | --- |
| Domestic violence | Intimate partner violence in front of children's, a parent stabbing another parent, mental violence between parents, physical abuse from a parent | 49 |
| Sexual or physical violence | Childhood sexual abuse, incest victimization, rape | 73 |
| Other crimes | Bank robbery, drug crime, burglary, stalking, arson | 10 |
| Experiences of terrorism, war, or mass murder | School shooting, peacekeeper in war countries, plane hijacking, bomb attack | 8 |
| Suicides or suicide attempts | Suicide of a close other (a variety of specified methods), one's own failed suicide attempt | 23 |
| Illness/death of a close other, or participant's own illness | Cancer, long-term illness of a child, death of a child during childbirth, sudden death, alcoholism of a loved one, cerebral infarction | 130 |
| Natural disasters | Storm, wildfire, tsunami, earthquake, mudslide, volcanic eruption | 25 |
| Fire accidents | A fire started by lightning, a fire caused by an intoxicated person, workplace building fire | 24 |
| Other severe accidents | Car crash, train accident, Estonian sinking, plane crash, drowning/near miss, work accident, serious fall, deer accident, poisoning | 93 |
| Other sorts of experiences | Mold disaster in the house, legal dispute with a family member, partnership problems, animal attack, bankruptcies or other financial disasters, workplace disputes, patient injuries, disappearance of a loved one, child custody | 59 |
|  |  | **Total:**<br>**494** |

More detailed information on the questions is available in **Tables 1‒5.**

The raw data was originally organized so that each participant had exactly one row in the data set. Trauma-related data was mainly represented so that each question, either structured or open, described above, was represented using two columns in the data set: first, a *trauma column* including the variable name of which encoded both year of the measurement and the type of traumatic experience for the structured questions (e.g., death of child) or trauma description for the open questions; and second, the reported year of the traumatic experience. From these questions and for all participants that had non-missing data in these columns, *trauma observations* were extracted into a new data set, the **trauma data set**. In that data set, each row thus referred to a trauma observation and, thus, included the following columns:

1. participant id,
2. trauma variable (e.g., death of mother),
3. measurement year,
4. year of the traumatic experience, and
5. trauma description (i.e., a free-text field of an open question).

This data set served as the basis for further data processing.

### 2.3. Further processing of the trauma variables

In 2007 and 2018, participants and their parents could utilize at most 3 or 4 free-text fields to describe their “significant life events” or “catastrophes”. Some participants, however, reported multiple experiences within a single free-text field (e.g., “mother’s death and father’s cancer”). In these cases, we converted each observations with such a description to two or more observations, with one new observation for each trauma. Additionally, some parents reported experiences that had occurred before the participant’s birth (e.g., a parent’s exposure to an accident in their childhood or a parent’s exposure to war bombing). **Traumatic experiences that had occurred before the participant’s birth were excluded**, as we were interested in participants’ lifetime traumatic experiences.

Next, we obtained **complementary information** in this way:

**1.** We calculated the participant’s **age at exposure** to each trauma observation.
**2.** In the context of accidents/injuries (reported between 1983 and 1992), we **cross-tabulated the years of inpatient hospital care** (a separate variable available in the YFS data set) with the years of accidents/injuries. If a participant had been in hospital care in the same year as the exposure to injury or accident, we assumed that the injury or accident had resulted in inpatient hospital care.
**3.** Few participants had not reported the year of a traumatic experience. Whenever possible, we **imputed the missing year using external sources**: for example, the year when the national disaster or ship accident mentioned in the free-text field had occurred.
**4.** In the baseline measurement (1980), the year of parents’ divorce was not reported. If possible, we deduced the year of divorce from the later follow-ups. If this was not possible, we recorded the divorce as having occurred in 1979.
**5.** Some parents reported traumatic experiences that had occurred to their “son” or “daughter” (without specifying which child). In these cases, we **examined familial information in the YFS data set** (e.g., number of children, sex, birth year) to determine whether the reported traumatic experience had occurred to a YFS participant and included the traumatic experience only if occurred to the participant with high certainty.

Furthermore, as expected, it became clear that some traumatic experiences had been reported multiple times in our data set, resulting in **duplicate observations that needed to be removed**. This occurred for two reasons: first, participants had reported some traumatic experiences multiple times across different follow-ups. Additionally, some traumatic experiences had been reported by both participants and their parents. For example, a fire at home building in 1977 had been reported as an accident by a parent (in the follow-up of 1983) and as a significant negative life event by the participant (in the follow-up of 2007). Or, a participant reported a severe injury/illness of a close other in 1989, the same year as the death of a close other or a severe motor vehicle accident, and these were assumed to refer to the same case. In order to make the data set free from duplicates, we conducted **a case-by-case investigation, reviewing the traumatic experiences reported for each participant**. Here, we marked any identified duplicates as such and used the resulting unique traumatic experiences in further processing.

Due to decades of follow-ups and multiple respondents (the participant and/or their parent), there were minor discrepancies in the reported years of certain traumatic experiences. For example, a motor vehicle accident could have been reported by a parent as occurring in 1989 and by the participant as occurring in 1990. As a main rule, we used the year given in **the follow-up closest to the reported experience** to obtain the latest available information. **Regarding divorce/separation**, some participants had reported divorce/separation in two slightly different years, presumably because a divorce process may take several years (from preliminary decision to final confirmation). It seemed that participants had initially (in the follow-up closest to the reported experience) reported the year of the preliminary decision and later (in a subsequent follow-up) the year of the finalized divorce. In these cases, we used the earlier reported year, as the most significant distress likely occurs soon after making the decision to divorce. As an exception, some participants reported two or three divorces *in a same follow-up*, occurring in years close to each other (e.g., in the 2007 follow-up, a participant reported two separate divorces that had occurred in 1999 and 2000). In these cases, we included all reported divorces as trauma observations.

**Regarding accidents/injuries**, there were minor discrepancies in reported years when a trauma was reported several times. When removing duplicates, we considered the reported exposure year to a traumatic experience, the follow-up year when the traumatic experience was reported, and type of the injury/accident. Specifically, we treated two reported accidents as referring to the same event if the ICD codes were identical or very similar, and if, at the second reporting point, the participant indicated that the accident had occurred prior to the latter measurement (e.g., reporting a ’car accident’ for 1981 in the 1983 follow-up, and a ’traffic accident’ for 1982 in the 1986 follow-up). In such cases, we used the exposure year reported in the former measurement. Additionally, some ICD codes referred to ’late effects’ of accidents (i.e., residual conditions resulting directly from a past injury). As our primary interest was in the original traumatic experiences, we excluded ’late effect’ entries if they were reported for the same year as another injury. However, late effects were retained if the participant had not previously reported the initial traumatic experience.

#### An additional variable of mental disorder as traumatic experiences

In free-response variables, some participants had reported the onset or treatment of a mental disorder as a “catastrophic experience” or significant life event. Since this may confound in analyses predicting mental disorder outcomes, we added a new variable **mental disorder** in the trauma data set, with value 1 for cases where participants’ mental disorder composed the trauma and 0 otherwise, to enable exclusion of these trauma observations. Importantly, onset of mental disorders of close others (e.g., a spouse’s psychosis) was coded as 0.

### 2.4. Development of the Traumatic Experience Severity Scale (TESS)

As can be expected in a population-based dataset, the participants had reported a large variety of different experiences (e.g., an ankle fracture, mother’s suicide, a fatal car crash killing two people, death of a cat). To be able to consider the variance of experiences in our statistical analyses, we decided to score the experiences. No single dimension seemed to adequately capture the variety of the experiences. Thus, we developed **the Traumatic Experience Severity Scale (TESS) with six subscales, each with 0, 1, or 2 points.** The TESS score is calculated as the sum of scores of the subscales.

The subscales were designed based on (a) previous research literature on traumatic experiences (evidence on which factors are relevant when predicting the risk of posttraumatic stress disorder (PTSD) or other posttraumatic outcomes), and (b) by considering what information is available in the YFS data set. A detailed description of the subscales can be found in **Table 6**. A summary of the subscales is as follows:

**1) Rarity.** This subscale is a rough evaluation of how common, normative, or predicted the experience was in the light of population statistics (e.g., a less rare divorce vs. a more rare bull attack) and/or normative life course (e.g., less rare grandparent’s death vs more rare child’s death). More rare experiences are given higher scores. This rarity subscale is included because the predictability of a traumatic experience modifies the intensity of the initial reaction and the (un)availability of peer support from others with similar experiences. According to population statistics, relatively common events include divorce/separation from a partner (5), the death of a parent when offspring was middle-aged (over 35 years of age) (6, 7), or the death of a spouse when the participant was over 50 years old (8). Unexpected experiences include, for example, onset of a rare disease (9, 10) or a sudden medical event (11).
**2) Same-household status.** This subscale evaluates whether the person to whom the experience happened was living in the same household, including the self (2 points), or not (0 points). Some YFS participants, for example, reported domestic violence occurring at a cousin’s home, while others reported it happening in their own home, making it necessary to distinguish between these contexts. Previous evidence indicates that the proximity of a traumatic experience matters: PTSD is more prevalent and severe among individuals directly exposed to traumatic events, compared to those who merely heard about the event (12). Furthermore, reactions appear to be stronger when the affected person resides in the same household: the death of a sibling or offspring seems to have more detrimental effects on siblings or parents when it occurs at a young age, while living together in the same household (13-15). Thus, in this study, same-household status was used as an indicator of how closely the traumatic experience affected the individual.
**3) Effects on everyday life.** This subscale is a rough evaluation of whether a traumatic experience caused a death or permanent disability, temporary harm, or at most minor harm. Higher scores refer to more severe effects on everyday life. This subscale is based on previous research showing that severity of injury (16) and post-traumatic functional impairments (17) are significant predictors of post-traumatic mental health conditions. Taken together, this subscale estimates whether the experience likely resulted in impairments in occupational, social, or familial life.
**4) Shamefulness.** This subscale evaluates whether an experience was likely to cause shame or stigmatization for the participant. Higher scores refer to higher expected shamefulness. The subscale is included because posttraumatic shame is shown to influence one’s sense of self, reduce the likelihood of discussing the experience with others, thereby limiting available social support, diminish help-seeking behavior, and hinder emotional processing of the event (18). Consequently, shame is also a crucial factor in the development of later psychopathology such as PTSD (19). Prior research has identified shame as particularly common in three types of experiences: (a) adverse sexual or reproductive events, such as rape (20), child abortion (21), or infertility (22), (b) certain externally visible neurological or somatic diseases, such as epilepsy (23) or dystonia (24), and (c) severe mental health conditions, including psychosis (25), or familial suicide (26).
**5) Human-made physical threat.** This subscale evaluates whether an experience involved physical threat to the participant caused by another person, i.e., whether someone could be held responsible for the experience intentionally (2 points) or accidentally (1 point) or not (0 points, e.g., a natural disaster or an onset of a somatic disease). This subscale is based on prior evidence demonstrating that human-made events are more likely to trigger psychiatric conditions than non-human-made events such as natural disasters (27). Overall, a great body of evidence has demonstrated that a perceived threat/danger from another person, including sense of horror and adrenergic arousal (e.g., being threatened by a gun, a penetrating trauma, or assault), is a strong predictor of PTSD (28, 29). Importantly, even accidents *unintentionally* caused by another person can increase the risk of posttraumatic mental health conditions (30). More specifically, individuals exposed to an intentional trauma have increasing rates of PTSD over time, while those exposed to non-intentional trauma have decreasing rates of PTSD (31).
**6) Danger to life or health.** This subscale evaluates whether the experience involved death or a life-threatening danger (2 points, regardless of whether the experience resulted in long-term harm) or not (0 points). Thus, the purpose of this subscale is to capture the psychological/emotional shock of a life- or health-threatening experience. For instance, a spouse’s near-death experience typically elicits a strong emotional response, even if the situation ended happily. This subscale is based on previous evidence showing that, over several decades, individuals have consistently rated life-threatening and health-related events as more distressing than e.g. economic hardships (32-34). Furthermore, a perceived sense of life endangerment has been linked to an increased risk of developing PTSD (35).

**Table 6.** Characteristics of the subscales with examples.

|  | Examples of scoring |  |  |
| --- | --- | --- | --- |
|  | 0 points | 1 point | 2 points |
| **Rarity**<br>How common, normative, or expected the experience was in the light of population statistics and/or normative life course? | Relatively expected or common experiences<br>▪ Divorce<br>▪ Breakdown in relationship with a close one<br>▪ Death of a grandparent<br>▪ A piece of property stolen<br>▪ Economic hardships<br>▪ Bullying in a workplace<br>▪ A mild storm or flood<br>▪ Miscarriage<br>▪ Death of spouse when being at age of > 50 years<br>▪ A comparatively common disease/disorder (e.g., a heart infarction, an anxiety period, asthma, a benign tumor) | Quite rare but not extraordinary experiences<br>▪ Sudden accidents of a relatively common nature (e.g., a car crash)<br>▪ Onset of a severe illness in adulthood<br>▪ A health condition that is comparatively mild but simultaneously in many family members (e.g., benign tumor in several family members)<br>▪ Infertility or unexcepted reproductive issues<br>▪ Disability pension or debt bondage<br>▪ Parent's death when the participant was < 35 years old (6, 7)<br>▪ Death of spouse when being < 50 years old (5)<br>▪ Death of a sibling/friend after reaching middle age (> 35 years)<br>▪ Emotional or physical violence targeted at adults | Very rare and unexpected experiences<br>▪ Rare and severe accidents (e.g., complete destruction of one's home in a fire, the sinking of the Estonia, the death of a whole family at once)<br>▪ Victim to an attempted manslaughter, murder, or terroristic crime<br>▪ Death of a parent/sibling when being < 18 years old<br>▪ Death or severe illness of one's own child (e.g., brain cancer, a severe congenital disease)<br>▪ Extreme sorts of natural disasters (e.g., exposure to a tsunami)<br>▪ The brutal death or suicide of a close person (e.g., family murder)<br>▪ Violent crimes targeted at children<br>▪ Exceptional sorts of lawsuits |
| **Same-household status**<br>Was the person to whom the experience happened living in the same household (incl. the self)? | The affected person was *not* likely living in the same household<br>▪ A grandmother<br>▪ An adult-aged offspring<br>▪ A colleague or friend<br>▪ A parent/sibling after the participant had reached adulthood<br>▪ A second-degree relative, such as a grandparent or aunt | This was scored as 0 or 2 | The affected person was likely living in the same household<br>▪ Participants themselves<br>▪ Participant's underage child<br>▪ Partner<br>▪ Note: Exceptionally also grandmother or another person if the participant reported to be living with him/her |
|  | 0 points | 1 point | 2 points |
| **Effects on everyday life** Did the experience cause permanent or long-term changes to everyday life, a temporary impairment, or at most minor/transient harm? | At most mild/transient effect on everyday life<br>▪ A mild illness with a good prognosis<br>▪ A minor injury that did not likely cause severe impairments<br>▪ Transient hardships in working life<br>▪ An eyewitness to an accident<br>▪ Exposure to a mild natural event without injuries (e.g., storm)<br>▪ Suspicion of illness that was ruled out<br>▪ A near-miss incident<br>▪ Parents' financial hardship after the participant has moved out of the home<br>▪ Death of a cat<br>▪ An illness or death of a second/third/fourth-degree relative (except for grandparents or grandchildren) | A temporary disruption to everyday life<br>▪ An illness or injury with severe but short-term restrictions to daily life (e.g., a heart infarction)<br>▪ An accident resulting in late effects but not inpatient hospital care<br>▪ Victimization to a comparatively mild crime (e.g., residential burglary)<br>▪ Death or severe illness of a parent/sibling during adulthood while living in a separate household<br>▪ Death of a friend<br>▪ Miscarriage<br>▪ Long-term unemployment<br>▪ A resolved relationship crisis<br>▪ Criminal death or suicide of a second/third/fourth-degree relative (e.g., a grandparent) | Long-term or stable effects on everyday life<br>▪ One's own severe illness (e.g., a poor-prognosis cancer)<br>▪ Severe accidents resulting in inpatient hospital care (e.g., an airplane crash, an attack of a big animal, a head-on collision with car)<br>▪ A severe illness of a person who is living in the same household (e.g., child's severe illness or disability)<br>▪ Divorce from spouse<br>▪ Parents' divorce or death (if being < 18 years old and likely living with parents)<br>▪ First-degree family member dies as a victim of crime<br>▪ Full fire at house<br>▪ Becoming homeless<br>▪ Exposure to domestic/physical violence |
|  | 0 points | 1 point | 2 points |
| **Shamefulness** Did the experience likely cause shame or stigmatization for the participant? | The experience was *not* likely especially shameful or stigmatizing<br>▪ A somatic illness/injury that usually is not stigmatizing (e.g., a cardiovascular event, a foot injury, a cancer)<br>▪ Adverse but quite common life events (e.g., divorce)<br>▪ Natural disasters<br>▪ Death of a close person with no known shameful reasons<br>▪ Victim to a financial crime or a mild crime (e.g., theft or residential burglary)<br>▪ Shameful types of experiences if the affected people are outside the family (e.g., violence in a cousin's family) | Experiences that commonly cause stigma or shame *but* are quite common in the population<br>▪ Economic or work-related setbacks of the individual or their parents that are partially public (e.g., debt restructuring, bankruptcy)<br>▪ Mental disorders with relatively good prognosis (e.g., depression)<br>▪ Relatively common issues related to sexual or reproductive health (e.g., infertility, miscarriage)<br>▪ An outwardly visible somatic illness with quite a good prognosis (e.g., epilepsy)<br>▪ A public work-related failure (e.g., an electoral defeat)<br>▪ A sexual or violent crime against a close person by someone outside the household (e.g., the rape of a sister) | Experiences that commonly cause stigma, shame, or delays in disclosure to others<br>▪ Domestic/sexual violence toward the self<br>▪ Severe mental disorders (e.g., psychosis, mania)<br>▪ A suicide or suicide attempt<br>▪ Rare sexual or reproductive conditions (e.g., a POF diagnosis, Klinefelter syndrome, chromosomal abnormality)<br>▪ Substance use disorders within the household (e.g., parent's alcoholism)<br>▪ Child custody removal<br>▪ Child's congenital disability or a severe long-term illness<br>▪ An offspring's involvement in criminal behavior |
|  | 0 points | 1 point | 2 points |
| **Human-made physical threat**<br>Did the participant experience an accidental or intentional *physical threat* from another person? | Experiences that did *not* involve a physical threat to the participant's life or health from another person<br>Any mental or somatic disease<br>Participant's own suicide attempt<br>Secondary experiences (the participant was not exposed to threat)<br>Natural disasters<br>Accidents that did not involve any other person (e.g., moose accident)<br>Divorce or marital crisis<br>Economic hardships<br>Death of another person without the self being at risk | Experiences that likely involved an *accidental* physical threat from another person<br>Accidents involving other people (e.g., a car crash, the sinking of the Estonia) | Experiences that likely involved an *intentional* physical threat from another person<br>Exposure to a terroristic crime<br>Arson in an apartment building<br>Victimization to physical/sexual violence<br>Domestic violence<br>Presence in a shooting incident<br>A parent's/spouse's disorder with aggressive behavior (e.g., antisociality, substance abuse)<br>Occupational threat incidents (e.g., a police officer being assaulted) |
| **Life- or health-threatening danger**<br>Did the experience involve death or a life-threatening danger? | The experience did *not* likely include a death or life-threatening danger<br>An economic hardship<br>A court proceeding<br>Divorce/separation<br>A minor accident<br>Infertility<br>A mild disease<br>A minor surgical operation (e.g., a knee surgery) | The experience likely included a death or life-threatening danger<br>Fire at house<br>A physical assault, such as robbery or rape<br>An earthquake or a tsunami<br>A threatening accident, such as a car crash, a ship accident, or a poisoning<br>The birth of a child with a severe illness | This was scored as 0 or 1 |

Examples of traumatic experiences at different levels of the TESS score are presented in **Table 7**.

**Table 7.** Examples of the TESS scores.

| Score | Typical examples |  |
| --- | --- | --- |
| 0–2 | ▪ Death of a grandparent<br>▪ Death of a close pet<br>▪ A minor accident (e.g., a fall or fracture)<br>▪ A minor natural incident (e.g., a storm without injuries) | ▪ A mild illness<br>▪ A minor surgical operation (e.g., knee)<br>▪ Employee negotiations at workplace<br>▪ Transient crisis with partner |
| 3–4 | ▪ A transient but significant change in social/occupational life (e.g., divorce, job loss)<br>▪ A traffic accident with moderate effects<br>▪ Death of a <u>second/third-degree</u> relative (e.g., a grandchild) or a friend<br>▪ Death of a parent in adulthood | ▪ A long-term illness with a relatively good prognosis (e.g., a back disease, infertility treatments)<br>▪ A severe illness of a close other living <u>in a separate household</u> (e.g., friend's or sibling's cancer)<br>▪ A quite rare but harmless natural event (e.g., volcanic eruption) |
| 5–6 | ▪ A life-threatening accident that resulted in inpatient hospital care (e.g., fire at home, traffic accident)<br>▪ Onset of a malign disease (e.g., cancer)<br>▪ Severe illness or death of a spouse/child<br>▪ Loss of pregnancy<br>▪ Suicide of a close other | ▪ Severe natural disasters (e.g., tsunami)<br>▪ Exceptional incidents <u>outside one's household</u> (e.g., the death of a close family at once)<br>▪ Emotional/physical violence from a partner in adulthood |
| ≥ 7 | ▪ Particularly tragical death/severe injury of one's child (e.g., victim to violence, a devastating illness)<br>▪ An extremely rare and stigmatizing condition (e.g., chromosomal abnormality)<br>▪ Extreme types of crimes (e.g., airplane hijacking, school shooting, bomb attack) | ▪ Target/victim of an attempted homicide/murder<br>▪ Brutal types of domestic violence (e.g., a parent shooting a child, a parent stabbing the other parent, incest)<br>▪ Brutal suicide of a close other |

## 3. Descriptive results about the trauma data

Our trauma data includes a total of 7769 traumatic experiences. The frequency distributions of traumatic experiences with respect to the TESS score and age at trauma are illustrated in **Figures 2 and 3**. In the TESS, the highest frequences were obtained below 5 points, but there were close to 1000 experiences belonging to the most severe category, i.e., at least 7 points (**Figure 2**). Traumatic experiences had occurred most typically at the age of 15-25 years, with the earliest traumas occurring during the birth year and the latest traumas occurring in middle age (**Figure 3**).

**Figure 2.**
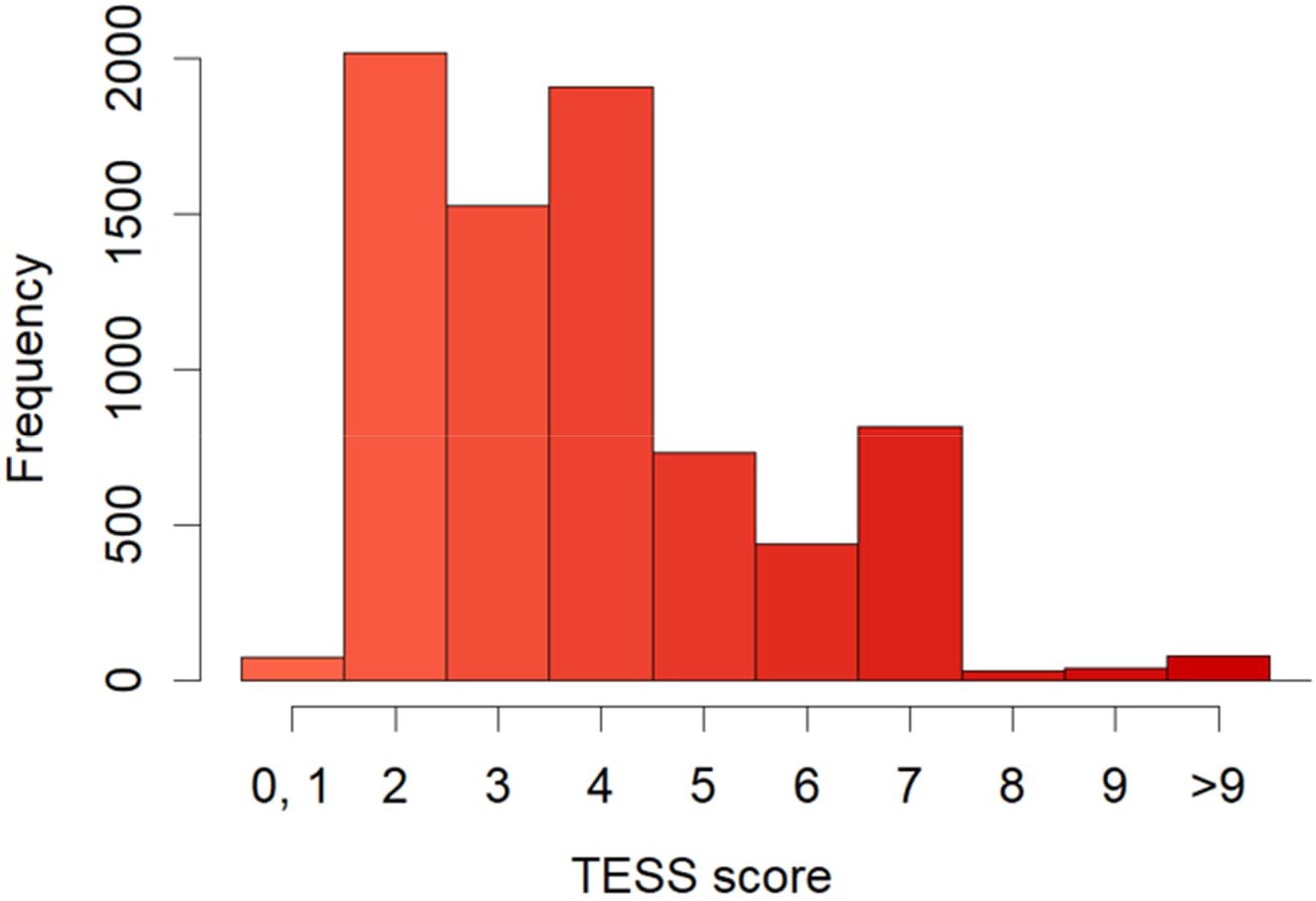
A histogram of the TESS scores.

**Figure 3.**
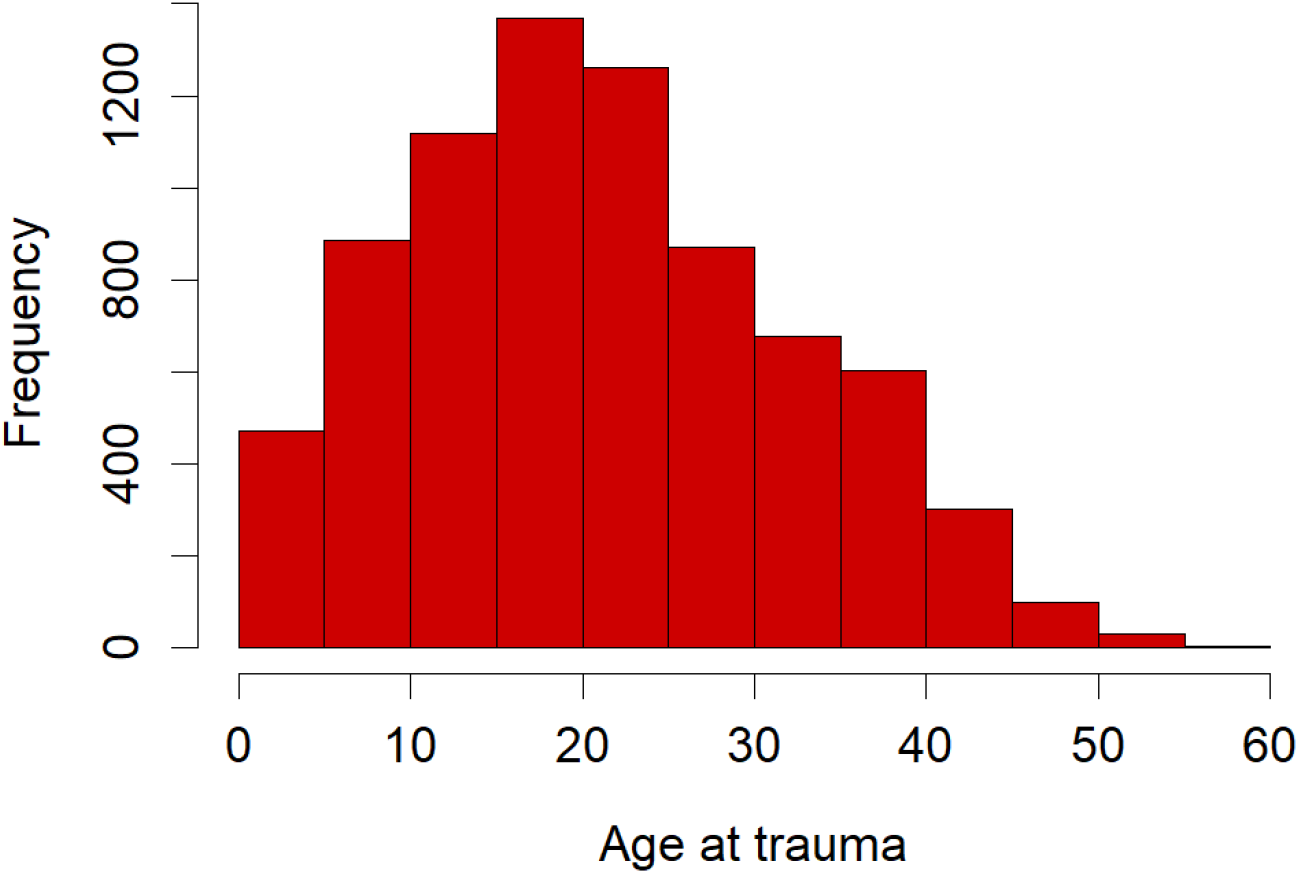
A histogram of the age at exposure to traumatic experiences.

## 4. Interleaving trauma data with longitudinal measurements

Taken together, the final trauma data set contained the following columns:

1. participant id,
2. trauma variable (e.g., death of mother),
3. measurement year,
4. year of the traumatic experience,
5. trauma description (i.e., a free-text field of an open question),
6. age at trauma
7. mental disorder,
8. rarity,
9. same-household status,
10. effects on everyday life,
11. life- or health-threatening danger,
12. shamefulness, and
13. human-made physical threat.

In this section, we describe how a data set (D) containing measurements of a variable, termed the *data variable* here, is interleaved with the trauma data set. Data set D is assumed to contain the columns participant id (ID), year and value.

To be able to account for the pretraumatic level of the dependent variables, each data set D of the dependent variables was joined with itself so that each pair of consecutive follow-ups formed a row in the combined data set D, with columns from both original copies of D. A row in the combined data set D* hence contains data of a follow-up *period* of a subject. In a combined data set, variables (*value* and *year*) related to the former measurement where subscripted with *x* and those related to the latter with *y*. The difference between follow-ups, i.e. *value_y_ – value_x_* was added to the data set as variable *diff*. In addition, the first and last follow-ups were included in the data set, with only the *y* and *x* subscripted variables (*value* and *year*) available, respectively.

The trauma data set T was used to augment the data sets D* as follows. If a trauma that occurred in year *t* was reported by the subject during that same year *t*, the trauma was *counted* to have occurred in year *t*. Otherwise the trauma was counted to have occurred in year *t* + 1: the idea is that if a trauma was reported in the same year as the data variable, the trauma likely already had an effect on the data variable, otherwise not. Let *t*′ denote the year when trauma was counted to have occurred. For each row *r* of D*, the traumas that were counted to have occurred in year *t*′ between *year_x_* (not included) and *year_y_* (included), ie, *year_x_ < t*′ ≤ *year_y_*, were considered; denote this set T*_r_*. For set T*_r_*, the following values were computed: (1) the maximum year *t*_max_ of occurrence of the traumas in T*_r_* was used to calculate: (2) *years since trauma = year_y_ – t*_max_ (3) the maximum value of each TESS subscale and mental disorder, (4) the number of traumas *n* in *T,* (5) indicator variables has trauma = 1 and no trauma = 0. These variables were then added to the data set D*. For rows r of D* with no associated traumas, the value 0 was assigned for these new variables, except *no trauma* = 1.

The final, interleaved data set combining trauma and longitudinal measurements was obtained as follows using D^+^ for each subject:

1. Starting from the first period, include it and all the consecutive periods such that no one of them has a trauma, if any such period exists
2. For the set of periods above, take the first of them in sequence (*a*) and every period b from the 3rd period onwards, add periods with values *year_x_* and *value_x_,* and *n* from *a*, columns *year _y_* and *value_y_* from from *b*, and *diff*, *years since trauma* and *n* computed as above. These rows give additional follow-ups without trauma.
3. Include all periods with trauma
4. For a period *a* (with trauma) and consecutive periods *b* (without trauma) after *a*, add periods with values *year_x_* and *value_x_,* and *n* from *a*, columns *year_y_* and *value_y_* from from *b*, and *diff*, *years since trauma* and *n* computed as above. These rows give additional follow-ups for a trauma (*a*) without interfering traumas.

## 5. Conclusions

We developed a new, comprehensive trauma dataset, covering a broad range of traumatic experiences from childhood to middle age over a 38-year follow-up. On the basis of the population-based YFS dataset, we also developed a new instrument – the Traumatic Experience Severity Scale (TESS) – to assess the severity of traumatic experiences at the population level. We hope this dataset can respond to many gaps of the previous trauma research. As the YFS dataset also includes decades of follow-up of psychological measures, cardiovascular data, epigenetic variables, and lifestyle factors, we believe that integrating this trauma data to the other measures provides exceptional possibilities for examining the longitudinal interplay between traumatic experiences and health.

## Data Availability

The Cardiovascular Risk in Young Finns (YFS) dataset comprises health-related participant data, and their use is therefore restricted under the regulations on professional secrecy (Act on the Openness of Government Activities, 612/1999) and on sensitive personal data (Personal Data Act, 523/1999, implementing the EU data protection directive 95/46/EC). Due to these legal restrictions, the data from this study cannot be stored in public repositories or otherwise made publicly available. However, data access may be permitted on a case by case basis upon request. Data sharing outside the group is done in collaboration with YFS group and requires a data-sharing agreement. Investigators can submit an expression of interest to the chairman of the publication committee (Prof. Mika Kähönen, Tampere University, Finland).

